# Feasibility of a novel non-invasive swab technique for serial whole-exome sequencing of cervical tumors during chemoradiation therapy

**DOI:** 10.1101/2022.02.23.22271256

**Authors:** Julianna K. Bronk, Chiraag Kapadia, Xiaogang Wu, Bhavana V. Chapman, Rui Wang, Tatiana V. Karpinets, Xingzhi Song, Andrew M. Futreal, Jianhua Zhang, Ann H. Klopp, Lauren E. Colbert

## Abstract

**Background:** Clinically relevant genetic predictors of radiation response for cervical cancer are understudied due to the morbidity of repeat invasive biopsies required to obtain genetic material. Thus, we aimed to develop a novel noninvasive cervical swab technique to collect tumor DNA with adequate throughput to perform whole-exome sequencing (WES) at serial time points over the course of chemoradiation therapy (CRT).

**Methods:** Cervical cancer tumor samples from patients undergoing chemoradiation were collected at baseline, at week 1, week 3, and at the completion of CRT (week 5) using a noninvasive swab-based biopsy technique. Swab samples were analyzed with whole-exome sequencing (WES) with mutation calling using a custom pipeline optimized for shallow whole-exome sequencing with low tumor purity. Tumor mutation changes over the course of treatment were profiled.

**Results:** 217 samples were collected and successfully sequenced for 70 patients. A total of 33 patients had a complete set of samples at all four time points. The mean mapping rate was 98% for all samples, and the mean target coverage was 180. Overall mutation frequency decreased during CRT with disease response but mapping rate and mean target coverage remained at >98% and >180 reads at week 5.

**Conclusion:** This study demonstrates the feasibility and application of a noninvasive swab-based technique for WES analysis to investigate dynamic tumor mutational changes during treatment and may be a valuable approach to identify novel genes which confer radiation resistance.

## Introduction

The global burden of cervical cancer is growing despite the development of HPV vaccines aimed at disease prevention^1^. In 2018, approximately 570,000 new cases of cervical cancer were diagnosed worldwide, resulting in more than 311,000 deaths^2^. High-risk human papillomaviruses (HPVs) are essential in cervical dysplasia and carcinogenesis and cause most cervical cancers^3^. Multimodality therapy is the standard of care for treating locally advanced disease and involves daily external beam radiation treatment, brachytherapy, and weekly chemotherapy ^4^. The rate of tumor regression during chemoradiotherapy (CRT) is variable and is strongly associated with survival^5,6^; however, predictive markers of radiation treatment sensitivity and resistance are currently unknown.

Whole-exome sequencing (WES) technology is a powerful tool that allows for comprehensive analysis of the frequency of somatic mutations, overall tumor mutational burden, and single nucleotide human exome variants (SNVs), which can lead to the identification of pathways that may be functionally significant in cancer outcomes^7^.

Previous comprehensive studies of cervical cancer genomics have relied on the analysis of untreated tumors, and none have identified predictors of radiation response^8,9^. This is mainly due to the challenges of repeated biopsy sampling over the course of the therapy, which are logistically challenging to acquire and entail significant patient morbidity. Nevertheless, cervical tumors are an ideal setting for the serial study of treatment response because tumors can be readily monitored by physical exam and are accessible for sampling through the course of chemoradiotherapy. Encouragingly, successful non-invasive swab-based sample acquisition of DNA has been demonstrated in the screening and diagnosis of precancerous and cancerous lesions in the oral cavity and detection of HPV-associated cutaneous lesions by PCR^10–13^.

This study hypothesizes that non-invasive swab-based sampling of cervical tumors can serve as a robust, noninvasive method to acquire tumor specimens for WES. Using swab-based biopsies collected prospectively in 70 patients undergoing CRT for newly diagnosed cervical cancer, we: (1) acquire tumor DNA samples adequate for WES in the majority of patients and (2) develop a custom pipeline optimized for shallow WES with low tumor purity (TP) to facilitate mutation calling.

## Materials and Methods

### Patient population and treatment characteristics

Patients were enrolled in an IRB-approved (2014-0543) multi-institutional prospective clinical trial at the University of Texas MD Anderson Cancer Center and the Harris Health System, Lyndon B. Johnson General Hospital Oncology Clinic (**Figure 1A, Table 1**). Inclusion criteria were newly diagnosed cervical cancer per the Federation of Gynecology and Obstetrics (FIGO) 2009 staging system, clinical stage IB1-IVA cancers, and visible, an exophytic tumor on speculum examination with planned definitive treatment of intact cervical cancer with external beam radiation therapy, cisplatin, and brachytherapy. Patients with any previous pelvic radiation therapy were excluded.

**Figure 1.**
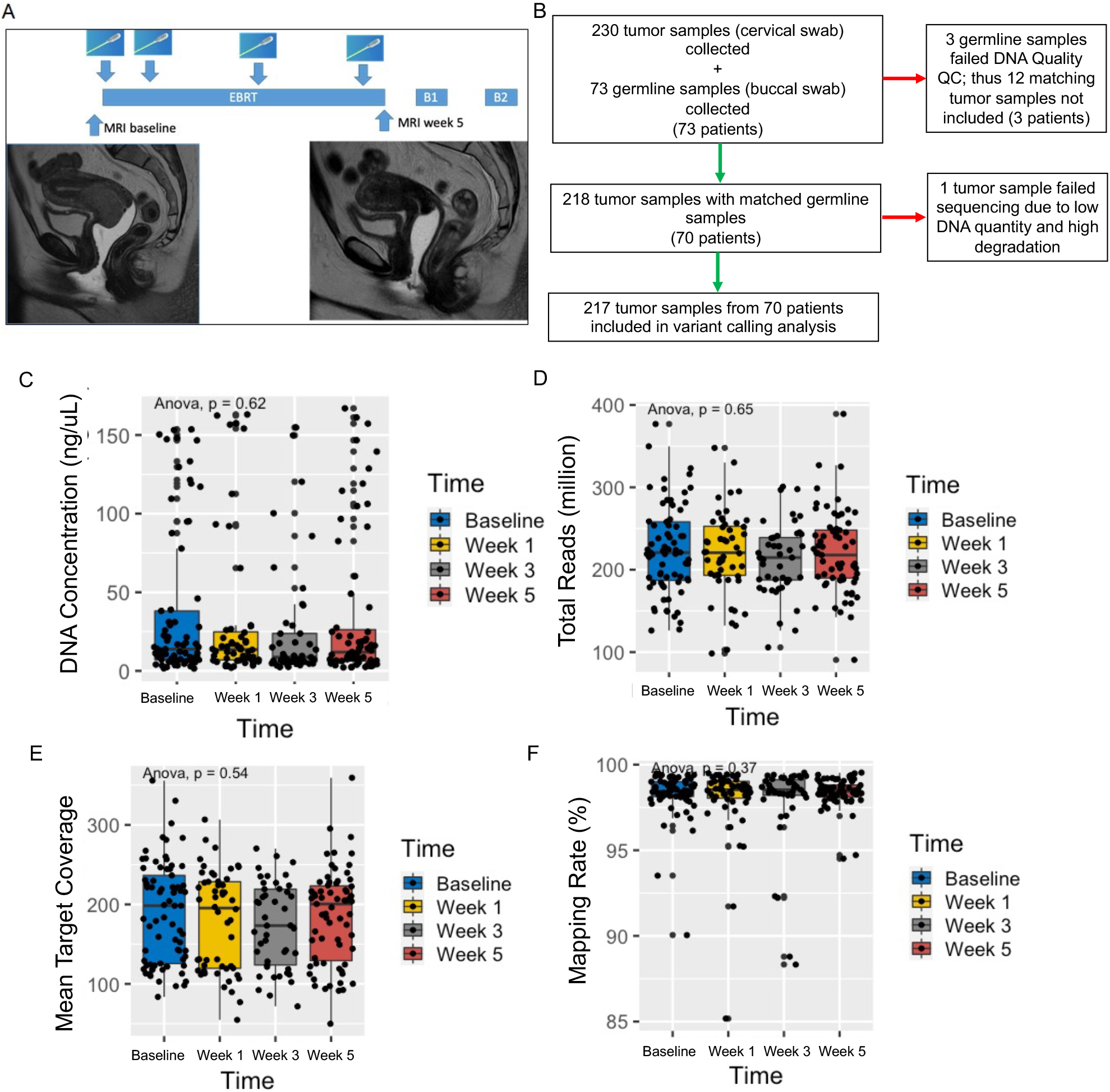
Overall Study Design and Quality Characteristics for all Samples by Time. (A) Patients with cervical cancer underwent five weeks of external beam radiation therapy (EBRT) followed by two brachytherapy treatments (B1 and B2), with swab samples collected at baseline, week 1, week 3, and week 5 of radiation therapy. (B) Of the 73 patients accrued on protocol, 70 patients with 218 total samples had DNA of adequate quantity and quality for sequencing. One tumor sample failed sequencing due to low quantity and high degradation and was not included in the analysis. (C) Total DNA concentration recovered did not differ by acquisition timepoint. Total reads (D), mean target coverage (E), and mapping rate (F) did not vary by timepoint.

**Table 1.** Patient Characteristics

| Characteristic | No. of patients | % |
| --- | --- | --- |
| **Age at diagnosis** |  |  |
| Median | 47 years |  |
| Range | 28-91 years |  |
| **Race/Ethnicity** |  |  |
| African American | 5 | 7% |
| American Indian/Alaskan Native | 2 | 3% |
| Asian | 2 | 3% |
| Hispanic/Latino | 31 | 44% |
| White/Caucasian | 30 | 43% |
| **Histology** |  |  |
| Serous | 1 | 1% |
| Adenocarcinoma | 9 | 13% |
| Squamous cell carcinoma | 60 | 86% |
| **2009 FIGO stage** |  |  |
| IA1 | 1 | 2% |
| IB1 | 4 | 6% |
| IB2 | 12 | 17% |
| IIA | 5 | 7% |
| IIB | 31 | 44% |
| IIIB | 12 | 17% |
| IVA | 5 | 7% |
| **Tumor grade** |  |  |
| I | 3 | 4% |
| II | 24 | 34% |
| III | 24 | 34% |
| Unknown | 19 | 27% |
| **Highest clinically involved nodal level on PET or CT** |  |  |
| Para-aortic | 4 | 6% |
| Common iliac | 12 | 17% |
| External iliac | 23 | 33% |
| Internal iliac | 6 | 9% |
| Node-negative | 22 | 31% |
| Unknown | 3 | 4% |
| **Max tumor dimension on MRI, cm** |  |  |
| Median | 5 |  |
| Range | 1.2-11.5 |  |
| Unknown | 2 |  |

Patients underwent standard-of-care pretreatment evaluation for disease staging, including tumor biopsy to confirm the diagnosis; pelvic magnetic resonance imaging (MRI) and positron emission tomography/computed tomography (PET/CT); and standard laboratory evaluations, including a complete blood cell count, measurement of electrolytes, and evaluation of renal and liver function. Patients received pelvic radiation therapy to a total dose of 40-45 Gy delivered in daily fractions of 1.8 to 2 Gy over 4 to 5 weeks. Thereafter, patients received intracavitary brachytherapy with pulsed-dose-rate or high-dose-rate treatments. According to standard institutional protocol, patients received cisplatin (40 mg/m^2^ weekly) during external beam radiation therapy. Patients underwent repeat MRI at the completion of external beam radiation therapy or at the time of brachytherapy, as indicated by the extent of disease.

### Sample collection and DNA extraction

Isohelix swabs (product # DSK-50 and XME-50, www.isohelix.com, UKSamples) were brushed against the viable cervical tumor several times by a clinician from the department of radiation oncology or gynecologic oncology at either The University of Texas MD Anderson Cancer Center or Lyndon B. Johnson General Hospital Oncology Clinic. The isohelix swab has a unique matrix design that yields one to five micrograms of high-quality DNA sufficient for sequencing applications from a single swab of the tumor surface^14^. Patients underwent swabbing at baseline, the end of week 1 (after five fractions), at the end of week 3 (after 10-15 fractions), and within a week before the first brachytherapy treatment or at the time of brachytherapy (week 5), for a total of four swabs during radiation therapy (**Figure 1A**). Additionally, 9 patients had swabs collected at the first follow up visit after treatment completion (week 12). In each swabbing session, attention was taken to obtain samples from the same general tumor region. Normal buccal samples were collected once at baseline to identify germline mutations present in individual patients. DNA was extracted from normal buccal and cervical cancer samples per Isohelix # DSK-50 manufacturer’s instructions.

### Whole-exome sequencing and mutational analysis

Illumina WES sequencing was performed on normal buccal control and cervical tumor DNA swab samples. Captured libraries were sequenced on a HiSeq 4000 series (Illumina Inc., San Diego, CA, USA) on a TruSeq version 3 Paired-end Flowcell according to manufacturer’s instructions at a cluster density between 700–1000K clusters/mm^2^. Sequencing was performed for 2 × 100 paired-end reads with a 7-nucleotide read for indexes using Cycle Sequencing version 3 reagents (Illumina). The average coverage achieved with the Roche Nimblegen probes was 180 reads (range 50-359) for cervical tumor samples.

Paired-end raw sequence reads in fastq format were aligned to the reference genome (human Hg19) using Burrows-Wheeler Aligner (BWA)^15^ with three mismatches with 2 in the first 40 seed regions for sequences less than 100 bp or using BWA-MEM with 31 bp seed length for sequences over 100 bp. The aligned BAM files were subjected to mark duplication, re-alignment, and re-calibration using Picard and the Genome Analysis Toolkit^16^ before any downstream analyses.

A custom computational pipeline was optimized for mutation calling (**Figure 2**). Based on the alignment results (BAM files) above, somatic mutations, including single-nucleotide variants (SNVs) and small insertions and deletions (INDELs), were obtained through merging variants from multiple somatic variant callers – MuTect, Pindel, GATK4 Mutect2, and Strelka2^17–20^. Common population variants reported in dbSNP138, 1000Genomes, ESP6500, and EXAC with >1% allele frequency were removed. The following mutation-filtering criteria were applied for calling somatic mutations: (i) sequencing depth ≥ 20 for tumor and ≥10 for normal, (ii) tumor variant allele frequency (VAF) ≥ 2%, and normal VAF < 2%, (iii) Evidence (number of somatic variant callers supported) ≥ 2. Associations between somatic mutations and disease progression/treatment response were analyzed and visualized using Maftools^21^. Tumor purity (TP), that is, the proportion of cancer cells in a tumor sample, was calculated from SNVs by the Tumor Purity Estimation (TPES) as well as from copy number profiles using Sequenza ^22,23^ (**Table S1**).

**Figure 2.**
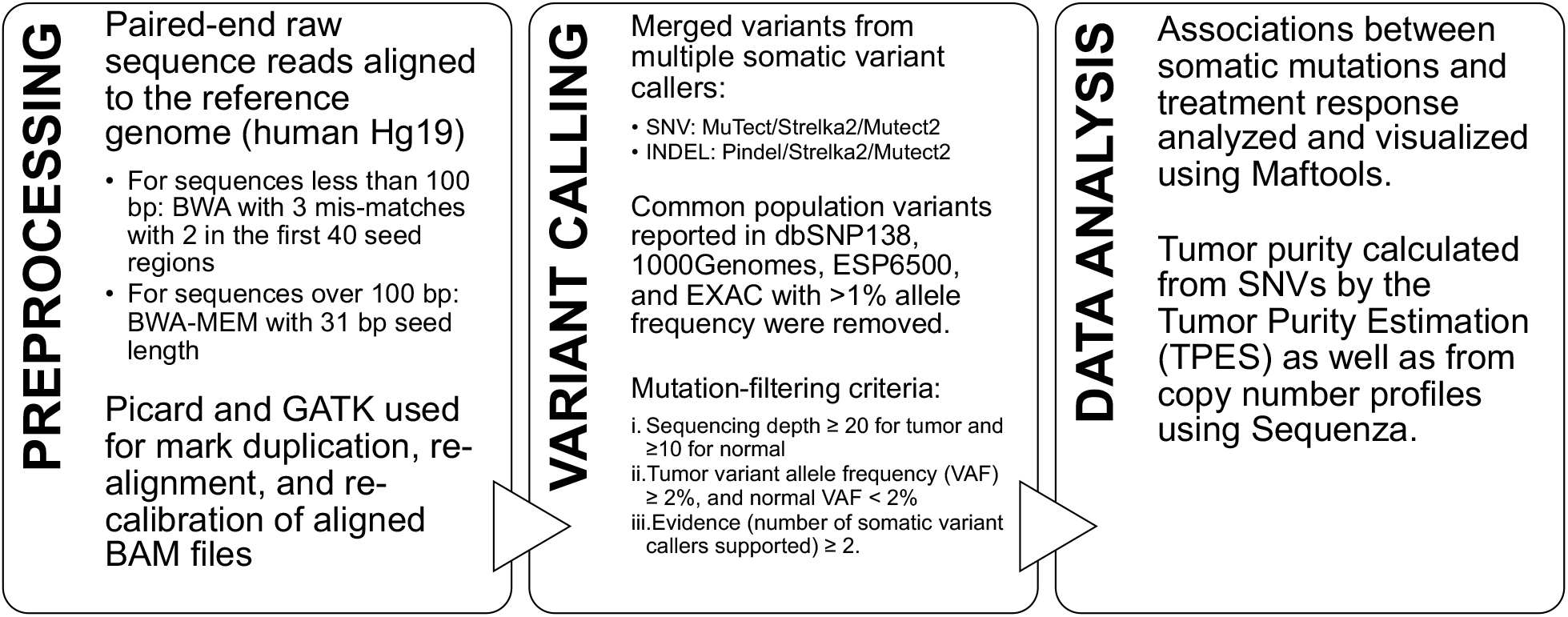
Computational Pipeline for Whole Exome Sequencing Data. Workflow depicting preprocessing, variant calling and data analysis tools and parameters implemented to analyze WES data acquired from tumor DNA collected by cervical swab.

## Results

### Patient and tumor characteristics

Clinicopathologic data are summarized in **Table 1**. Overall, 48 patients (68%) had advanced disease (stage IIB or greater), and most had squamous cell carcinoma with tumor grade II or higher. The median tumor size (based on the short axis diameter on pretreatment MRI) was 5cm (range 1.2-11.5 cm). Forty-five patients had positive pelvic or para-aortic lymph nodes on PET or CT.

### DNA quality and sequencing characteristics of collected samples

Two hundred thirty total tumor samples were collected (**Figure 1**). For 3 patients, matching normal (buccal) samples failed either DNA quality or quantity quality check (QC), and thus 12 paired tumor samples were excluded. Two hundred eighteen total tumor samples and 70 normal germline samples were sequenced. Of these, 161 (74%) had both optimal DNA quality and quantity QC, 54 (25%) had suboptimal quantity, but adequate quality and 2 (1%) had optimal quantity but low-quality DNA. 1 sample failed sequencing due to both suboptimal DNA quality and quantity; thus, 217 total tumor samples from 70 patients were included in the analysis (**Figure 1B**). Median DNA concentration per sample was 12 ng/uL (range 1.5-167 ng/uL). The mean total reads per sample was 216 million (+/-50), and mean mapping rate was 98.15% (+/-1.84) (**Table 2**). Total DNA concentration was not different by timepoint (p=0.62), nor were total reads (p=0.65), mapping rate (p=0.37), or mean target coverage (p=0.54) (**Figure 1C-F**). No significant differences were detected by sequencing batch.

**Table 2.** Quality Characteristics for Samples by Timepoint

|  | Baseline (N = 67) | Week 1 (N = 49) | Week 3 (N = 42) | Week 5+ (N = 60) |
| --- | --- | --- | --- | --- |
| **DNA concentration (ng/uL)** |  |  |  |  |
| Mean/SD | 37.55 ± 48.40 | 32.37 ± 47.38 | 25.99 ± 38.11 | 34.22 ± 47.24 |
| Median | 13.80 | 13.55 | 8.70 | 11.90 |
| **Total Reads (million)** |  |  |  |  |
| Mean/SD | 224.46 ± 53.86 | 219.62 ± 51.31 | 212.29 ± 42.84 | 220.58 ± 49.42 |
| Median | 220.75 | 220.64 | 214.74 | 217.78 |
| **Mapping Rate (%)** |  |  |  |  |
| Mean/SD | 98.29 ± 1.33 | 98.02 ± 2.24 | 97.86 ± 2.51 | 98.38 ± 0.86 |
| Median | 98.45 | 98.53 | 98.53 | 98.44 |
| **Mean Target Coverage (per sample)** |  |  |  |  |
| Mean/SD | 189.07 ± 63.46 | 179.29 ± 62.41 | 173.22 ± 53.73 | 184.87 ± 59.70 |
| Median | 198.360 | 195.385 | 173.330 | 200.510 |
| **Insertions/ Deletions (Count)** |  |  |  |  |
| Mean/SD | 42 ± 20 | 42 ± 30.5 | 39 ± 29 | 36 ± 21 |
| Median | 38 | 36 | 35 | 32 |
| **Substitutions (Count)** |  |  |  |  |
| Mean/SD | 296 ± 402 | 353 ± 529 | 281 ± 510 | 124 ± 241 |
| Median | 168 | 164 | 137 | 55 |
| **Sequenza Tumor Purity** |  |  |  |  |
| Mean/SD | 0.28 ± 0.20 | 0.27 ± 0.20 | 0.23 ± 0.17 | 0.22 ± 0.15 |
| Median | 0.195 | 0.180 | 0.150 | 0.185 |
| **TPES Tumor Purity** |  |  |  |  |
| Mean/SD | 0.61 ± 0.22 | 0.60 ± 0.19 | 0.61 ± 0.25 | 0.56 ± 0.23 |
| Median | 0.630 | 0.590 | 0.615 | 0.580 |

### Mutation characteristics for all samples

Sixty-seven patients had samples analyzed at baseline, 49 at week 1, 42 at week 3, and 60 at week 5 or later. 33 patients had samples analyzed at all four time points. Mean number of substitutions, insertions and deletions over time was 337 (range 50-2,857) at baseline, 394 (range 48-2,281) at week 1, 321 (range 57-2,223) at week 3 and 160 (range 38-1,843) at week 5 or greater (**Table 2, Table S1**).

Median TP of all samples as calculated by the Sequenza algorithm decreased from baseline (median 0.195 [range 0.10-0.99]) to week 5 (median 0.185 [range 0.10-0.98]; p=0.046; **Table 2, Table S1**). Median TP of all samples as calculated by the TPES algorithm at baseline was 0.61 (range 0.17-0.99) and 0.55 (range 0.12-0.97) at week 5 (p=0.097; **Table 2, Table S1**).

Across analyzed samples (n=217), the top 5 most common alterations were in PIK3CA, FBXW7, FNDX1, KMT2D, and CSMD3 (**Figure 3**). For all 33 patients with samples available at all 4 time points (n=132 samples), the top 10 most common alterations were in PIK3CA (17%), FBXW7 (13%), KMT2D (11%), LRP1B (11%), RYR2 (11%), MUC16 (10%), CSMD3 (8%), EP300 (8%), PCLO (8%), and KMT2C (8%) (**Figure 4**). Of these, the remaining detectable alterations at week 5 were in FBXW7, LRP1B, and RYR2. From the top 50 alterations, other genes with alterations remaining at week 5 included CGREF1, CAMSAP1, DOCK11, LRP2, CRTAC1, KIF2, LTPB1, STK11, SUFU, GPR98, AGGF1, AMER1, CLTCL1, and ENC1.

**Figure 3.**
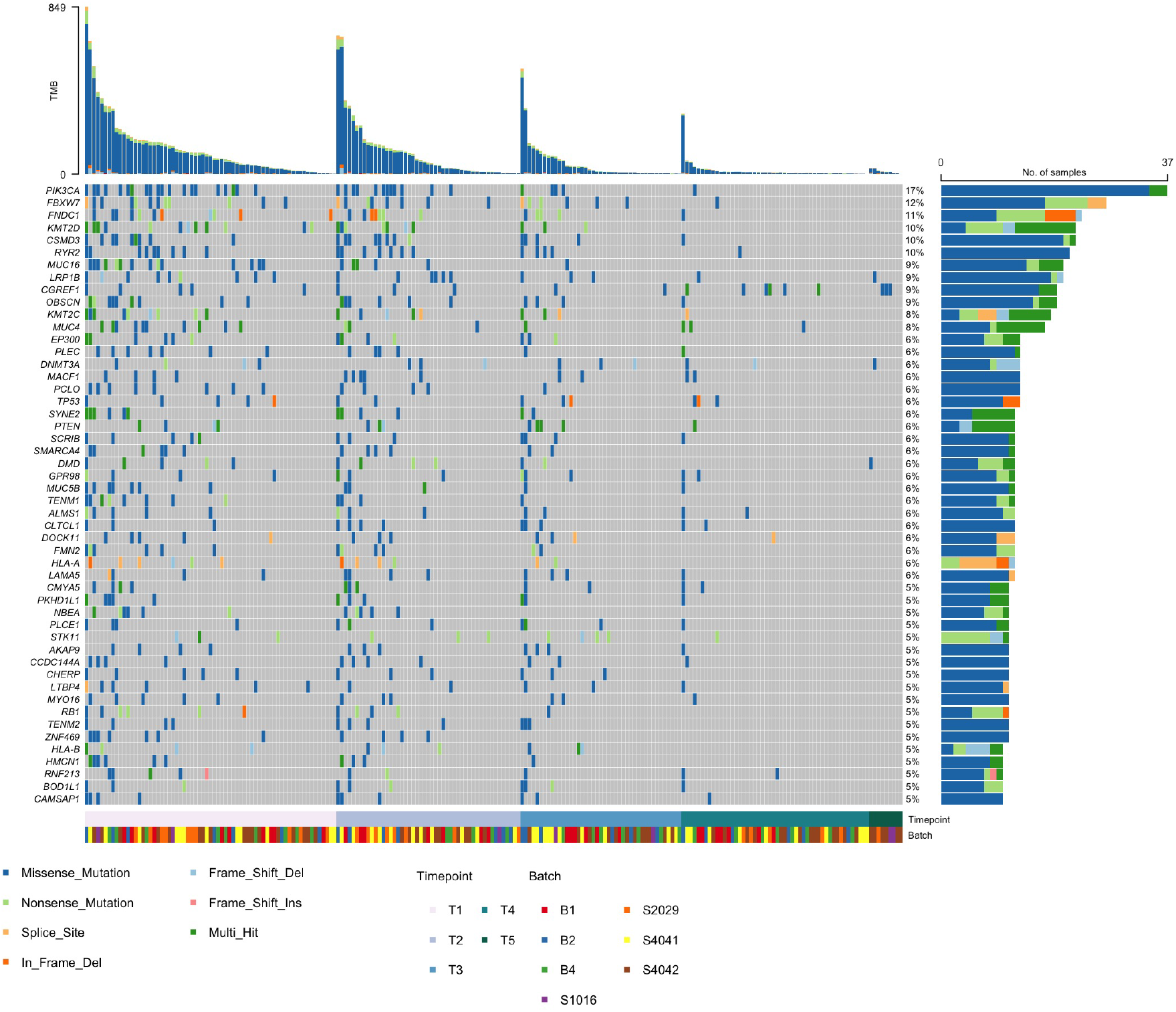
Top 50 Gene alterations over time for 70 patients with paired normals. Heat map displaying the top 50 genes ranked by occurrence for 70 patients (217 samples) grouped by timepoint collection during chemoradiation.

**Figure 4.**
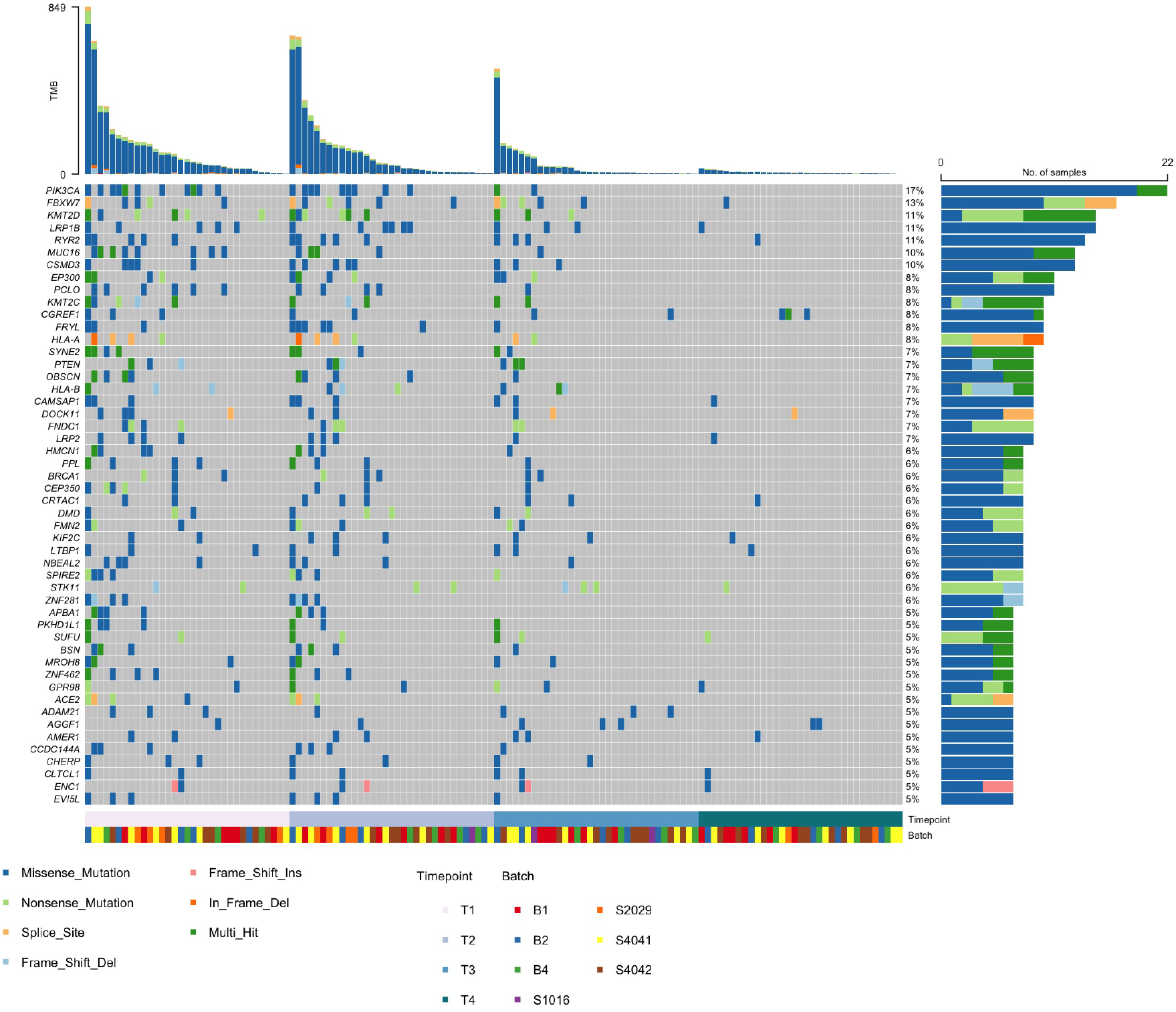
Top 50 Gene alterations over time for 33 patients with all four-time points. Heat map displaying the top 50 genes ranked by occurrence for 33 patients (132 samples) grouped by timepoint collection during chemoradiation.

## Discussion

To our knowledge, this is the first study describing the use of non-invasive swab-based cervical tumor sample collection and performance of serial WES over the course of CRT. TP analysis confirmed the presence of tumor cells captured and sequenced at timepoints throughout the course of treatment, and gene alterations present at baseline and persistent through the course of treatment were identified. We anticipate that this methodology will allow for comprehensive characterization of changes in the mutational landscape of cervical cancers in patients undergoing treatment.

Past studies that relied on serial biopsies have been limited by logistical challenges, concerns about patient discomfort, and complications from traditional biopsies, including bleeding and infection. In a report by Weidhaas et al., gene expression profiling was performed on tissue biopsies collected from 13 patients pre- and mid-treatment and investigators identified a 7 gene signature that predicted improved local control^24^. In a similar study of patients with locally advanced cervical cancer undergoing CRT, investigators acquired biopsies pretreatment and at week 3 of CRT and performed RNA-sequencing on 20 matched pairs. They found that patients who succumbed to disease at the time of their report had enrichment of gene expression from mitotic pathways and increased retention of HPV E6/E7 gene expression at week 3 of CRT, which may promote treatment resistance^25^. No analysis of somatic mutations was performed in either of these studies. Our novel, non-invasive technique allows DNA to be collected from cervical swabs, reducing obstacles to serial biopsy collections. Furthermore, while previous studies have recognized gene expression signatures associated with long-term patient outcomes, this is the first study to report findings from WES of cervical tumor samples collected longitudinally over the course of CRT.

As expected, TP estimates for swab acquired samples were lower than previously reported estimates from the Cancer Genome Atlas (TCGA) analysis in biopsy samples^26^ and decreased with disease response. While several computational methods exist to infer TP, these methods differ in the types of genomic information used, such as gene expression, SCNA, and somatic mutations^27,28^. To reduce the systematic bias of genomic investigation of samples containing both tumor and non-neoplastic tissue, TP levels are often considered during analysis to deconvolute contaminant contributions from the tumor microenvironment field^29–31^. Thus, we evaluated two purity estimation tools with differing prediction mechanisms to better understand the cellular heterogeneity within our swab acquired samples. The Sequenza tool was developed for both exome and whole-genome deep sequencing of tumor DNA where average depth ratio in tumor versus normal samples and allele frequency is used to estimate for cellularity and ploidy^23^. Investigators found that Sequenza correctly detected ploidy in samples with as low as 30% tumor content. TPES predicts TP from variant allelic fraction distribution of SNVs to more accurately predict TP when tumor genomes are copy-number neutral or euploid^22^. This tool was validated on WES data from TCGA tumor samples and enabled TP estimation in samples that failed TP prediction algorithms dependent on somatic copy-number alterations. TPES estimates were enriched in samples with low genomic burden, while SCNA-based tools similar to Sequenza were more proficient with high genomic burden cases suggesting a complementary role for these two tools in the analysis of tumor WES data.

While the clinical significance of individual mutations was not the focus of the present study, studies are underway to identify signatures and mutations associated with radiation sensitivity and resistance among women with differential responses to CRT for cervical cancer. For this future work, we hypothesize that mutations that survive the initial weeks of radiation treatment may be clinically relevant drivers of radiation resistance, and we plan to focus on characterizing the clonal architecture of residual tumors to identify more granular molecular signatures predictive of treatment response. This will permit future investigations of treatment escalation or de-escalation in appropriate populations.

There are several limitations to this study that must be addressed. First, tumor samples were not available at all four time points for 37 patients due to the inability to procure samples. Second, long-term follow-up data is not available for this cohort at this time. An additional limitation of the current study is the absence of tumor biopsy samples with which to compare sequencing output to swab-acquired DNA. A previous study examining epigenome-wide DNA methylation compared sample acquisition by biopsy and isohelix brush swab for cancers in the oral cavity found no significant difference in DNA yield between tissue and brush samples and matched tissue. Isohelix brush swabs had an excellent correlation in the oral cavity^10^. In this prior study, mapping efficacy was over 90% for swab-acquired DNA. Investigators successfully identified potential prognostic and predictive biomarkers for malignant lesions in the oral cavity with high sensitivity and specificity using a comparable isohelix swab design as was used here^10^. While isohelix brushes have been designed and marketed for DNA acquisition of the buccal mucosa, we were able to acquire sufficient quality and quantity DNA for WES in all but one tumor sample with collected samples by isohelix brushings from cervical mucosa. This method could rapidly be applied to the study of other gynecologic, head and neck, anorectal, and skin malignancies to establish biomarker predictors for treatment response and identify potential drivers of treatment resistance, which may be undetectable from analyses of only the initial pretreatment biopsy.

## Conclusion

In conclusion, this work provides proof of concept that a noninvasive, swab-based biopsy technique can be utilized to serially sample tumors for in-depth sequencing analysis. This novel methodology can be added to the translational research armamentarium to interrogate tumor genetics and eventually tailor cancer-directed therapies.

## Data Availability

All data produced in the present study are available upon reasonable request to the authors

## Supplementary Materials

**Table S1**. Sample and sequencing quality control metrics and tumor purity estimates for each tumor sample.

